# Mental health service activity during COVID-19 lockdown: South London and Maudsley data on working age community and home treatment team services and mortality from February to mid-May 2020

**DOI:** 10.1101/2020.06.13.20130419

**Authors:** Robert Stewart, Evangelia Martin, Matthew Broadbent

## Abstract

The lockdown and social distancing policy response to the COVID-19 pandemic in the UK has a potentially important impact on provision of mental healthcare; however, there has been relatively little quantification of this. Taking advantage of the Clinical Record Interactive Search (CRIS) data resource with 24-hourly updates of electronic mental health records data, this paper describes daily caseloads and contact numbers (face-to-face and virtual) for home treatment teams (HTTs) and working age adult community mental health teams (CMHTs) from 1^st^ February to 15^th^ May 2020 at the South London and Maudsley NHS Trust (SLaM), a large mental health service provider for 1.2m residents in south London. In addition daily deaths are described for all current and previous SLaM service users over this period and the same dates in 2019. In summary, comparing periods before and after 16^th^ March 2020 the CMHT sector showed relatively stable caseloads and total contact numbers, but a substantial shift from face-to-face to virtual contacts, while HTTs showed the same changeover but reductions in caseloads and total contacts (although potentially an activity rise again during May). Number of deaths for the two months between 16^th^ March and 15^th^ May were 2.4-fold higher in 2020 than 2019, with 958 excess deaths.

## Background

The COVID-19 pandemic may have a profound impact on health services: i) because of the direct effects of the virus itself; ii) because of social distancing policies and their psychological impact; iii) because of the impact of social distancing policies on delivery of mental and physical healthcare. Mental healthcare faces a range of challenges including the heightened vulnerability of its patient populations (e.g. through cardiovascular and respiratory disorders), already-reduced life-expectancies (1), and frequently described problems accessing healthcare (2;3). In addition, services have had to be radically reconfigured to cope with suspected or confirmed COVID-19 infections in inpatient and outpatient settings, staff sickness or self-isolation, the need to minimise face-to-face contacts, and the need to accommodate increasing pressures on acute medical care from cases of viral pneumonia. These in turn are accompanied by the as yet unknown impacts of social distancing on already isolated or otherwise vulnerable populations, and of challenged national economies on already impoverished and disadvantaged communities. There is therefore a pressing need for information in the public domain (4).

Little has been reported to date on the mental healthcare impact of the COVID-19 pandemic. Taking advantage of a unique mental healthcare data research platform that receives 24- hourly updates from its source electronic records, we sought to describe daily activity in key services from February to mid-May 2020. At the time of writing it is envisaged that a series of open-access reports will be generated covering different elements of mental health service activity, and that this will be kept updated for as long as a single site’s data are deemed useful.

## Methods

The Biomedical Research Centre (BRC) Case Register at the South London and Maudsley NHS Foundation Trust (SLaM) has been described previously (5;6). SLaM serves a geographic catchment of four south London boroughs (Croydon, Lambeth, Lewisham, Southwark) with a population of around 1.2 million residents, and has used a fully electronic health record (EHR) across all its services since 2006. SLaM’s BRC Case Register was set up in 2008, providing researcher access to de-identified data from SLaM’s EHR via the Clinical Record Interactive Search (CRIS) platform and within a robust security model and governance framework (7). CRIS has been extensively developed over the last 10 years with a range of external data linkages and natural language processing resources (6). Of relevance to the work presented here, CRIS is updated from SLaM’s EHR every 24 hours and thus provides relatively ‘real-time’ data. SLaM’s EHR is itself immediately updated every time an entry is made, which include date-stamped fields indicating patient contacts (‘events’) and those indicating acceptance of a referral or a discharge from a given service (or SLaM care more generally). Mortality in the complete EHR (i.e. all SLaM patients with records, past or present) is ascertained weekly through automated checks of National Health Service (NHS) numbers (a unique identifier used in all UK health services) against a national spine. CRIS has supported over 200 peer reviewed publications to date. CRIS has received approval as a data source for secondary analyses (Oxford Research Ethics Committee C, reference 18/SC/0372).

Activity and caseload data were extracted via CRIS and enumerated for every day from 1^st^ February 2020 to 15^th^ May 2020. The focus of this report is on community mental health teams (CMHTs) for working age adults, and home treatment teams (HTTs), as well as overall mortality. SLaM provides separate community mental health services to children and adolescents, working age adults, and older adults, and working age CMHTs were a focus for this report. CMHTs included specialist teams for rehabilitation, complex needs, and eating disorders, but excluded specialist services for addictions, learning disability, perinatal mental health, disorder-specific outpatient clinics (e.g. post-traumatic stress disorder, chronic fatigue syndrome), dedicated psychological therapy services, forensic services, and any other national/tertiary services. HTTs comprised services to working age and older adults providing alternatives to inpatient care with the facility to provide at least daily assessments.

For CMHTs and HTTs, daily caseloads were calculated by ascertaining patients who were receiving active care from the service on a given day, based on the date a referral to that service was recorded as accepted to the point a discharge was made from that service. Daily contact numbers were ascertained from recorded ‘events’ (i.e. standard case note entries) for that service and were divided into the following groups according to structured compulsory meta-data fields for that event in the EHR: i) face-to-face contacts attended; ii) virtual contacts attended (by email, fax, mail, phone, online, or video link); iii) total contacts, as the sum of these two. Finally, mortality data (number of deaths for all patients with SLaM records) were extracted both for the period in question and for the same dates in 2019 as a comparison period (for this reason, data were omitted for 29^th^ Feb 2020). Data extractions were carried out on 3^rd^ June 2020.

Descriptive data were generated for the above parameters and displayed graphically. Weekend days and national holidays were omitted for CMHT contacts. For comparisons between time periods, lockdown was defined as commencing on 16^th^ March 2020, the date this policy was announced by the UK government. Mean (SD) activity levels were described before and after this date and percentage changes quantified. Mortality data were quantified for the two months from 16^th^ March to 15^th^ May and both the difference and the ratio calculated for numbers of deaths in 2020 and 2019.

## Results

Working age CMHT daily contacts are displayed in Figure 1 and daily caseloads are displayed in Figure 2. Mean (SD) total daily contacts were 702 (59) prior to 16^th^ March and 599 (139) subsequently (a 14.6% reduction); these were 548 (62) and 219 (70) respectively for face-to-face contacts (a 60.0% reduction), and were 154 (17) and 380 (97) for virtual contacts (a 147% increase). Mean (SD) daily caseloads were 8729 (24) before 16^th^ March and 8539 (124) subsequently (a 2.1% reduction).

**Figure 1.**
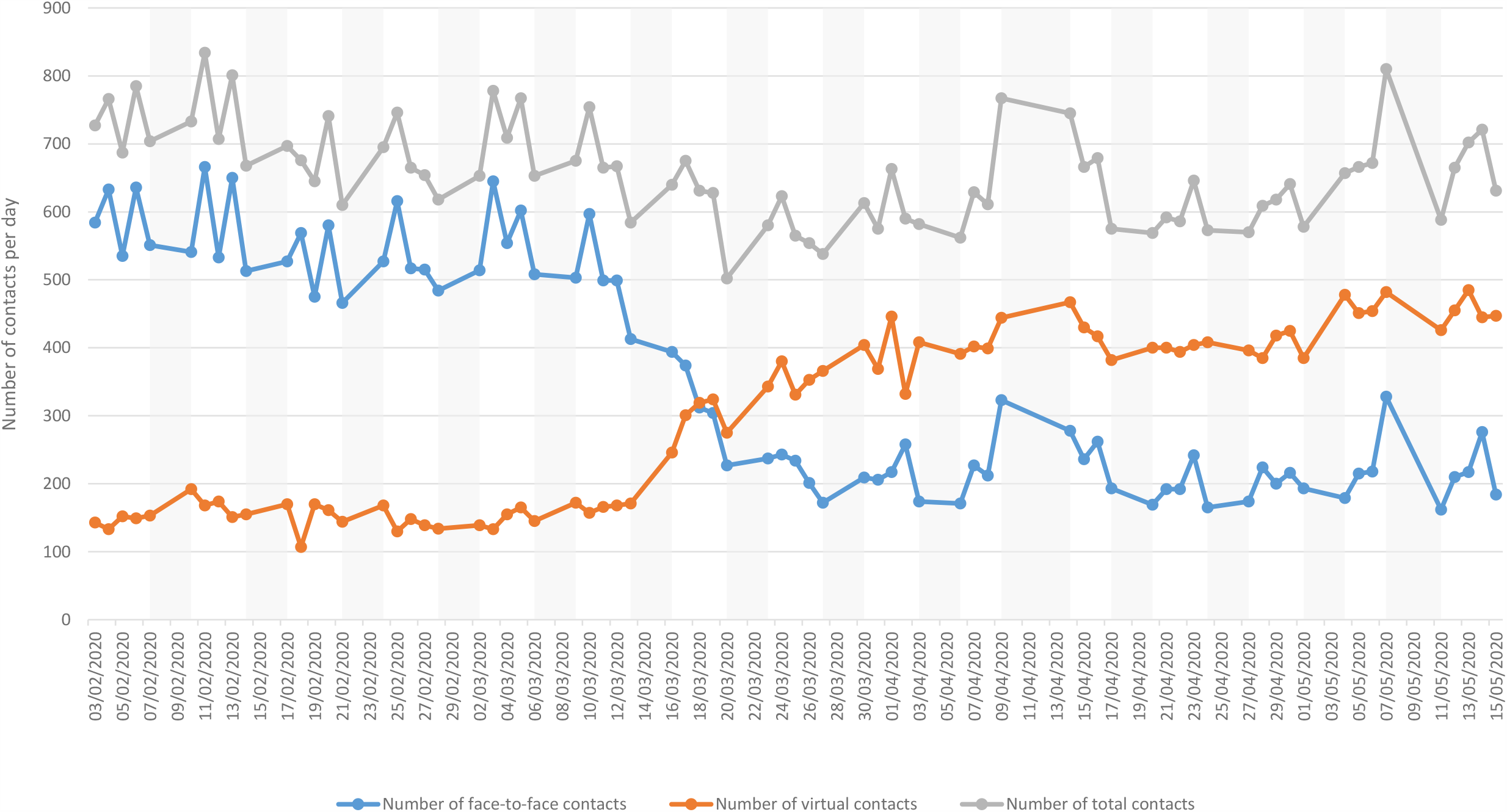
Working age adult Community Mental Health Team contacts (daily; Monday-Friday; February to mid-May 2020)

**Figure 2.**
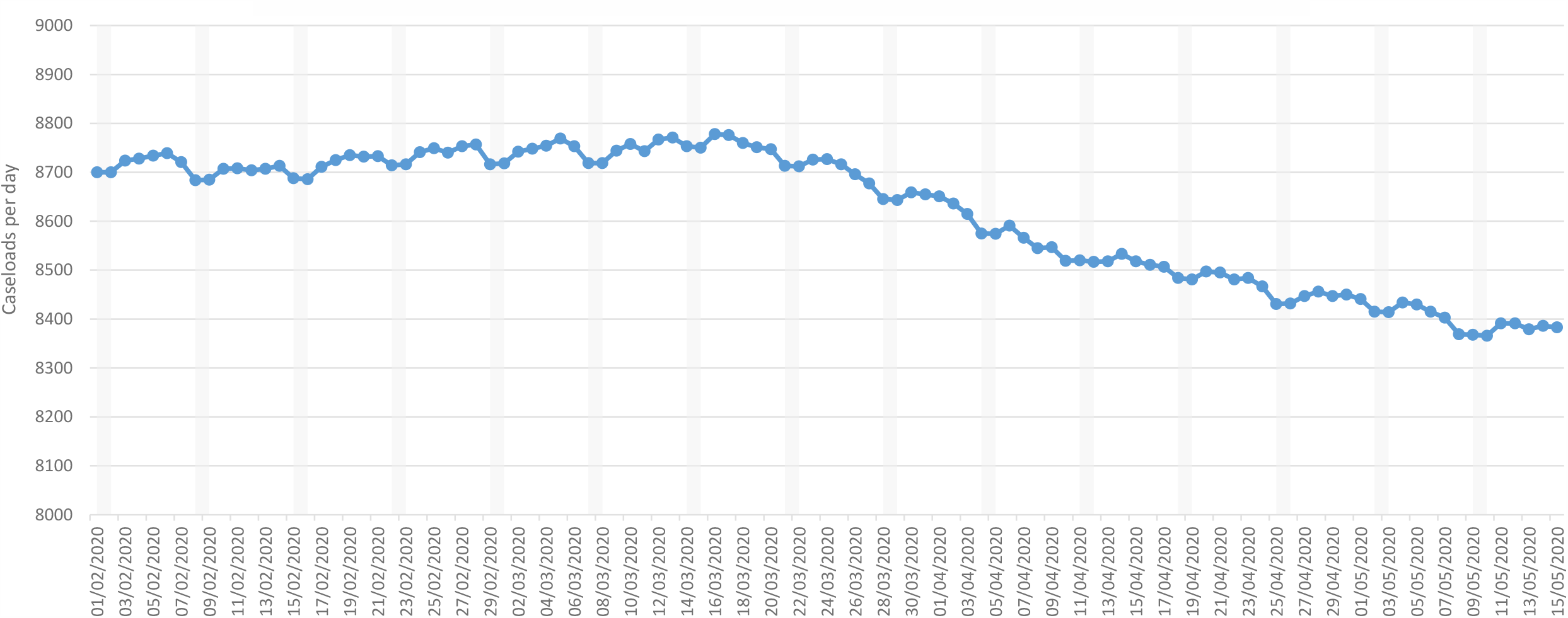
Working age adult Community Mental Health Team active caseloads (daily; February to mid-May 2020)

HTT daily contacts and caseloads are displayed in Figure 3. Comparing the period before 16^th^ March to that between 16^th^ March and 15^th^ May, mean (SD) face-to-face contacts decreased 50.2% from 135.2 (26.8) to 67.4 (18.0), mean virtual contacts increased 102.7% from 26.7 (7.3) to 54.2 (14.8), mean total contacts reduced 24.9% from 161.9 (30.7) to 121.5 (26.1), and mean daily caseload decreased 26.4% from 221.8 (8.5) to 163.3 (20.0). In a comparison of the period from 16^th^ March to 30^th^ April and that from 1^st^ to 15^th^ May, mean HTT daily caseloads had increased 12.2% from 158.6 (19.2) to 177.9 (15.1).

**Figure 3.**
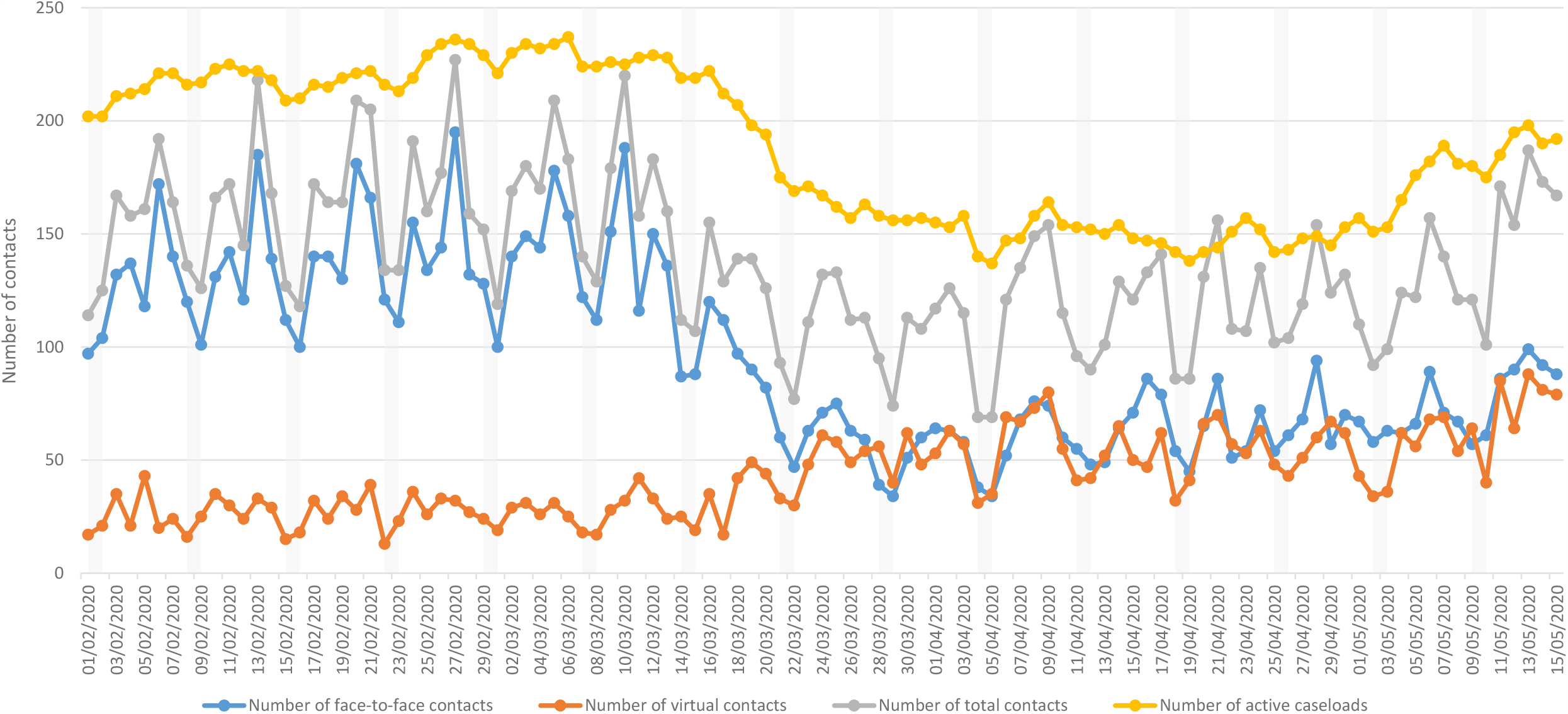
Home Treatment Team community contacts (daily; February to mid-May 2020)

Daily deaths for 2020 and the equivalent dates in 2019 are displayed in Figure 4. From 16^th^ March to 15^th^ May 2020 there were 1665 deaths, compared to 707 for the same period in 2019: a difference of 958 and a ratio of 2.36.

**Figure 4.**
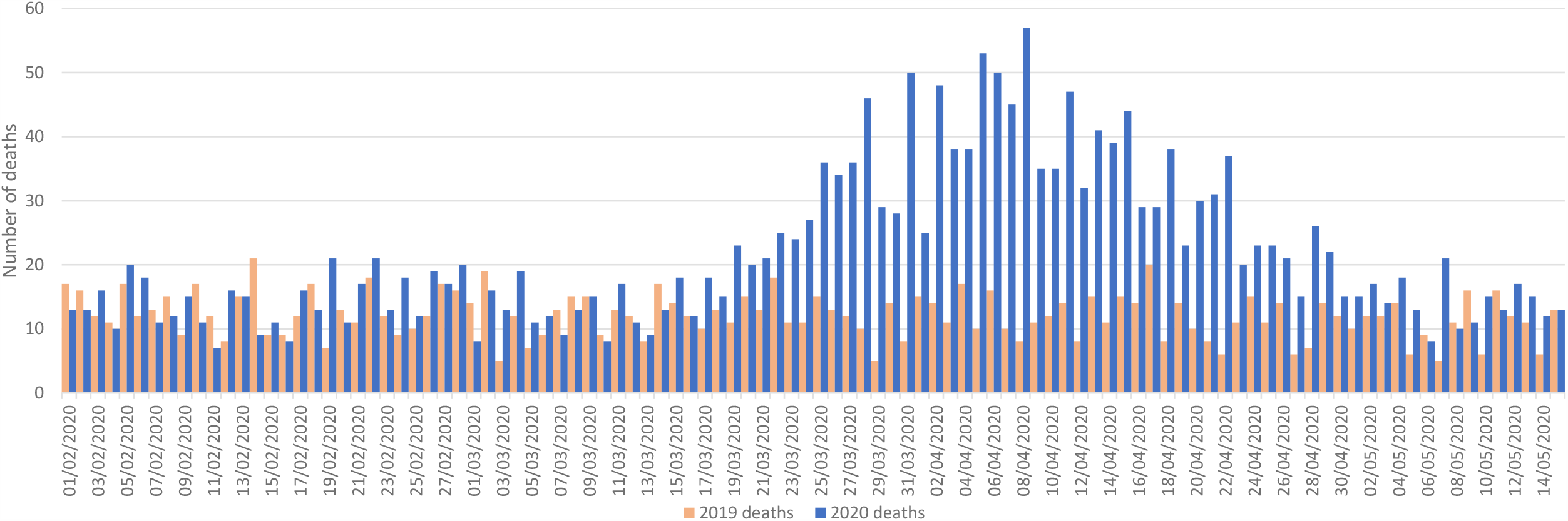
Total number of deaths in past/current SLaM patients (daily; February to mid-May 2020 with same-day comparisons from 2019)

## Discussion

Comparing periods before and after the 16^th^ March 2020 as the commencement of lockdown policy, working age CMHT caseloads and total contacts had sustained slight reductions (2.1% and 14.6% respectively) with more sizeable changes from face-to-face towards virtual contacts. HTT contacts showed a similar change but a larger total reduction (24.9%), and caseloads reduced 26.4% overall, although with some subsequent increase from 1^st^ May.

Daily deaths in past and current SLaM service users appeared to have returned to 2019 levels by mid-May, although having resulted in 958 excess deaths over the preceding two months since lockdown, 2.4 times higher than 2019 numbers.

Considering limitations, it is important to bear in mind that the data are derived from a single site. Because complete data are being provided for that site with no hypothetical source population intended, calculation of confidence intervals was not felt to be appropriate for the descriptive data provided in this report; applicability to other mental healthcare providers cannot therefore be inferred and would need specific investigation. Profiles of services and catchment morbidity are also likely to vary. While the service activity data are likely to be correctly represented from the source record, delays in national registrations might result in an underestimate of more recent deaths. Classification of service types (in this case into HTT/CMHT groupings) is inevitably approximate and likely to vary between mental health service providers.

## Data Availability

Data are available on request from the corresponding author.

